# *KLF3* and *PAX6* are candidate driver genes in late-stage, MSI-hypermutated endometrioid endometrial carcinomas

**DOI:** 10.1101/2021.04.26.21256125

**Authors:** Meghan L. Rudd, Nancy F. Hansen, Xiaolu Zhang, Mary Ellen Urick, Suiyuan Zhang, Maria J. Merino, National Institutes of Health Intramural Sequencing Center Comparative Sequencing Program, James C. Mullikin, Lawrence C. Brody, Daphne W. Bell

**Affiliations:** Cancer Genetics and Comparative Genomics Branch, National Human Genome Research Institute, National Institutes of Health, Bethesda, Maryland, United States of America; Computational and Statistical Genomics Branch, National Human Genome Research Institute, National Institutes of Health, Bethesda, Maryland, United States of America; Laboratory of Pathology, Center for Cancer Research, National Cancer Institute, National Institutes of Health, Bethesda, Maryland, United States of America; NIH Intramural Sequencing Center, National Human Genome Research Institute, National Institutes of Health, Rockville, Maryland, United States of America; Medical Genomics and Metabolic Genetics Branch, National Human Genome Research Institute, National Institutes of Health, Bethesda, Maryland, United States of America

## Abstract

Endometrioid endometrial carcinomas (EECs) are the most common histological subtype of uterine cancer. Late-stage disease is an adverse prognosticator for EEC. The purpose of this study was to analyze EEC exome mutation data to identify late-stage-specific statistically significantly mutated genes (SMGs), which represent candidate driver genes potentially associated with disease progression. We exome sequenced 15 late-stage (stage III or IV) non-ultramutated EECs and paired non-tumor DNAs; somatic variants were called using Strelka, Shimmer, Somatic Sniper and MuTect. Additionally, somatic mutation calls were extracted from The Cancer Genome Atlas (TCGA) data for 66 late-stage and 270 early-stage (stage I or II) non-ultramutated EECs. MutSigCV (v1.4) was used to annotate SMGs in the two late-stage cohorts and to derive p-values for all mutated genes in the early-stage cohort. To test whether late-stage SMGs are statistically significantly mutated in early-stage tumors, q-values for late-stage SMGs were re-calculated from the MutSigCV (v1.4) early-stage p-values, adjusting for the number of late-stage SMGs tested. We identified 14 SMGs in the combined late-stage EEC cohorts. When the 14 late-stage SMGs were examined in the TCGA early-stage data, only *KLF3* and *PAX6* failed to reach significance as early-stage SMGs, despite the inclusion of enough early-stage cases to ensure adequate statistical power. Within TCGA, nonsynonymous mutations in *KLF3* and *PAX6* were, respectively, exclusive or nearly exclusive to the microsatellite instability (MSI)-hypermutated molecular subgroup and were dominated by insertions-deletions at homopolymer tracts. In conclusion, our findings are hypothesis-generating and suggest that *KLF3* and *PAX6*, which encode transcription factors, are MSI target genes and late-stage-specific SMGs in EEC.

## Introduction

Endometrial carcinoma (EC) exacts a significant toll on women’s health. It resulted in 89,929 deaths globally in 2018 [1], and is projected to cause 12,940 deaths within the United States in 2021 [2]. Importantly, EC incidence is increasing annually in the US and many other countries [3]. This phenomenon is likely partly due to increasing rates of obesity [4], a well-recognized epidemiological risk factor for endometrioid endometrial carcinomas (EECs) that make up 75%-80% of all newly diagnosed endometrial tumors. EECs most often present as low-grade, early-stage (FIGO (International Federation of Obstetricians and Gynecologists) stage I or II) tumors, that are confined within the uterus [5]. Five-year survival rates for patients with low-grade, early-stage disease are high because surgery is often curative for this patient population, due to the limited extent of disease [5]. In contrast, patients with late-stage EEC have relatively poor outcomes [6], despite more aggressive treatment approaches of surgery with adjuvant chemotherapy or radiotherapy [7-9]. Thus, increasing tumor stage is an adverse prognosticator for EEC that is used in the clinical setting, as are high tumor grade (Grade 3; G3), and extent of lymphovascular space invasion [10]. The prognostic utility of molecular classification, according to *POLE*, microsatellite instability (MSI), and *TP53*/p53 status, is an area of active exploration originating from The Cancer Genome Atlas (TCGA) discovery that EECs can be subclassified into four molecular subgroups associated with distinct clinical outcomes [11](and reviewed in [12]).

TCGA’s initial comprehensive molecular characterization of primary endometrial carcinomas included exome sequencing of 200 EECs [11]; an expanded analysis that included 188 additional EECs was subsequently reported [13]. These studies confirmed prior findings that EEC exhibits high frequencies of somatic alterations resulting in activation of the PI3-kinase pathway, the RAS-RAF-MEK-ERK pathway, and the WNT/β-catenin pathway, frequent mutations in *ARID1A* (BAF250A) tumor suppressor, and mismatch repair (MMR) defects resulting in MSI [11, 13-15]. Moreover, many additional “significantly mutated genes” (SMGs), which represent candidate pathogenic driver genes, were annotated in EECs by TCGA using statistical approaches [11].

Given the dynamic nature of tumor genomes during disease initiation and progression, it is conceivable that the repertoire of pathogenic driver genes may differ in late-stage compared to early-stage EEC. However, the annotation of candidate driver genes (i.e., SMGs) in primary EEC exomes by TCGA, was performed in a stage-agnostic manner [11, 13]. An improved understanding of the molecular etiology of late-stage EEC may provide novel insights into disease pathogenesis and progression. The aim of this study was to delineate SMGs in late-stage EEC exomes, and to determine whether these genes are also significantly mutated in early-stage disease. To this end, we exome sequenced 15 “in-house” late-stage EECs (National Human Genome Research Institute (NHGRI) cohort) and reanalyzed somatic mutation calls from 66 late-stage and 270 early-stage non-ultramutated EECs within TCGA. Collectively, we identified 14 SMGs in 81 late-stage tumors. *KLF3* (Krüppel-like factor 3) and *PAX6* (Paired box 6), which encode transcription factors, were SMGs in late-stage tumors, but were not statistically significantly mutated in early-stage tumors. All *KLF3* mutations, and almost all *PAX6* mutations, were in the MSI-hypermutated EEC subgroup; within this subgroup, *KLF3* and *PAX6* mutations were more frequent in late-stage than early-stage tumors. The mutation spectrum of both genes included recurrent insertions-deletions (indels) at homopolymer tracts, consistent with strand slippage resulting from MMR defects, and suggesting that *PAX6* and *KLF3* are likely MSI target genes.

## Materials and Methods

### NHGRI clinical specimens

For 15 cases in the NHGRI cohort, anonymized, fresh-frozen endometrioid endometrial tumors and matched non-tumor (normal) samples were obtained from the Cooperative Human Tissue Network (CHTN) (**S1 Table**). The National Institutes of Health Office of Human Subjects Research Protections determined that this research was not human subject research, per the Common Rule (45 CFR 46). For each tumor sample, an H&E stained section was reviewed by an experienced gynecologic pathologist to identify regions containing ≥ 70% neoplastic cellularity; accompanying surgical pathology reports were retrospectively evaluated by the same gynecologic pathologist to annotate tumor stage using the FIGO (International Federation of Gynecology and Obstetrics) 2009 classification (**S1 Table**).

### Genomic DNA preparation and next-generation sequencing

Genomic DNA extraction, identity testing and MSI analysis of tumor and normal samples in the NHGRI cohort were performed as previously described [16]. DNA was purified by phenol-chloroform extraction prior to library preparation. DNA libraries were prepared using the SeqCap EZ Exome + UTR capture kit (Roche) and sequenced with the Illumina HiSeq 2000 platform (Illumina).

### Alignment and variant calling

Short sequence reads from NHGRI cohort exomes were aligned to the hg19 human reference sequence using NovoAlign version 2.08.02 (University of California at Santa Cruz). Four somatic mutation detection algorithms, Strelka [17], Shimmer [18], SomaticSniper [19], and MuTect [20], were used to call potential somatic variants. Insertions and deletions (indels) were identified by Shimmer and Strelka, while single nucleotide variants (SNVs) were identified by all four somatic algorithms. Strelka workflow version 1.0.14 (https://doi.org/10.1093/bioinformatics/bts271) was run with default parameters. Shimmer version 0.2 (https://github.com/nhansen/shimmer) was run with –min_som_reads=6 and -- minqual=20 [18]. SomaticSniper version 1.0.5 was run with options -Q 40 -G -L, followed by the “standard somatic detection filters” described in Larsen et al [19]. MuTect version 1.1.5 was run with default parameters, and data were then filtered to include only calls designated as “KEEP” in the program’s output [20]. Following analysis with each algorithm, a VarSifter-formatted file was generated containing the somatic variant allele frequencies observed in each tumor and matched normal sample for every called variant [21]. ANNOVAR (downloaded on August 12, 2014) was used to annotate all variants using the UCSC “known genes” gene structures [22].

### Variant filtering

Coding, splicing, and non-coding (intronic, 3’ or 5’ untranslated region (UTR), and 1kb upstream of the transcription start or downstream of the transcription end site) somatic variant calls in the NHGRI cohort were displayed using VarSifter [21]. A minimum of 14 reads covering a site in the tumor and 8 in the normal were required for mutation calling [23, 24]; potential germline variants (those with a variant allele frequency (VAF) of greater than 3% in matched normal samples) were excluded. Coding and splice-site single nucleotide variants (SNVs) were annotated against dbSNP Build 135 and nonpathogenic single nucleotide polymorphisms (SNPs) with a minor allele frequency (MAF) greater than 5% were excluded. Indel variants that were present in dbSNP Build 135 were excluded without further evaluation of MAF. SNVs called by all four algorithms and indels called by either Strelka or Shimmer were retained and further annotated against GENCODE hg19 using Oncotator (v1.5.3.0) (http://www.broadinstitute.org/oncotator) [25]; noncoding variants, those with a variant classification of UTR, Flank, lincRNA, RNA, Intron, or De novo start were excluded.

### TCGA data analysis

A subset of TCGA Uterine Corpus Endometrial Carcinoma (UCEC) somatic mutation data (TCGA UCEC PanCancer Atlas [13]) was extracted from the MC3 Public MAF file (mc3.v0.2.8.PUBLIC.maf.gz, https://gdc.cancer.gov/about-data/publications/mc3-2017) [26].

Briefly, the MC3 Public MAF file was filtered to include somatic variants from 336 EECs from the MSI-hypermutated (n=141), copy number-low/MSS (n=140) or copy number-high (n=55) molecular subgroups; variants from EECs within the ultramutated-POLE molecular subgroup or those without a molecular subgroup assignment were excluded (**S2 Table**). Molecular subtype annotation for each sample was obtained from the cBioPortal for Cancer Genomics [27, 28].

Variants with a PASS, WGA, or Native_WGA_mix designation as described by [26] were retained and further filtered to include SNVs called by MuTect and Indels called by Indelocator [13]. The final set of selected variants was annotated against GENCODE hg19 using Oncotator (v1.5.3.0) (http://www.broadinstitute.org/oncotator) [25]; noncoding variants, those with a variant classification of UTR, Flank, lincRNA, RNA, Intron, or De novo start were excluded. Additional clinicopathologic information for each tumor, including histology, stage, and grade, was obtained from Berger et al [13], and the cBioPortal for Cancer Genomics (URL: https://www.cbioportal.org/) [27, 28] (**S2 Table**). Early-stage tumors were defined herein as stage I or II; late-stage tumors were defined as stage III or IV.

### Annotation of statistically significantly mutated genes

Statistically significantly mutated genes (SMGs) were annotated using MutSigCV (v1.4). Briefly, MutSigCV (v1.4) was run on the NIH high-performance computing Biowulf cluster (http://hpc.nih.gov) using the coverage, covariate, and mutation type dictionary files provided by the Broad Institute. Filtered somatic variants for each data set were annotated against GENCODE hg19 using Oncotator (http://www.broadinstitute.org/oncotator) [25], noncoding variants were excluded in accordance with a published approach [29], and the resulting coding mutation annotation format (maf) files were uploaded to the Biowulf cluster. Somatically mutated genes with a false discovery rate (q-value) ≤0.10 were defined as SMGs in accordance with a published approach [30].

### Power analysis

MutSigCV’s statistical power to detect SMGs was estimated using the binomial model described in [30]. Briefly, the probability of obtaining a p-value <= 0.1/14 (for 14 tests) was calculated assuming a background mutation rate of *p*_0_ = 1 − (1 − *µf*_*g*_)^3/4*L*^, where µ is the background mutation rate, and *f*_*g*_*=3*.*9* and *L=1500* are the 90^th^ percentile gene-specific mutation rate factor and gene length, respectively. We also assumed a signal mutation rate of *p*_1_ = *p*_0_ + *r*(1 – *m*), where *r* is the frequency of non-silent mutations in tumor samples and *m=0*.*1* is the mis-detection rate. Power estimates were performed and plotted for a range of mutation rates and frequencies (**S1 Figure**) using an R script available at https://github.com/nhansen/LateStageEECs.

### Determining whether late-stage SMGs are statistically significantly mutated in early-stage tumors

MutSigCV (v1.4) was run as described above on the set of filtered somatic variants from the 270 early-stage EECs to obtain p-values for all mutated genes. For all genes annotated as SMGs in late-stage tumors, q-values were re-calculated from the MutSigCV (v1.4) p-values assigned to the early-stage data, adjusting for 14 tests (reflecting the total number of SMGs identified in late-stage tumors).

### *In silico* prediction of functional consequences for somatic variants

MutationAssessor [31], PROVEAN (Protein Variation Effect Analyzer) [32], SIFT (Sorting Intolerant From Tolerant) [33], and PolyPhen-2 (Polymorphism Phenotyping v2) [34], were used to predict the effects of missense mutations on protein function. For each algorithm, the following descriptors were considered as impacting protein function: “high” (MutationAssessor), “deleterious” (PROVEAN), “damaging” (SIFT), and “probably-damaging” (PolyPhen-2).

Agreement across at least three of the four prediction methods was required to assign an overall determination of “functional impact” for a missense mutation.

### Survival analyses

We utilized the cBioPortal for Cancer Genomics (https://www.cbioportal.org/) to query the relationship between SMG mutation status and survival (overall-, disease-free-, progression-free-, and disease-specific-survival) stratifying cases by stage (all stages, early-stage, late-stage) and molecular subgroup (MSI-hypermutated, CN-low, CN-high, all non-ultramutated), and applying a Bonferroni correction (number of SMGs multiplied by 48 (3 stage subgroups/4 molecular subgroups/4 survival subgroups)) to account for multiple testing.

## Results

### Identification of SMGs among late-stage EECs

For the NHGRI late-stage cohort (n=15), the average depth of coverage within regions targeted by the capture kit for tumor and normal samples was 67.2x and 65.5x, respectively; 90.87% of targeted bases for each tumor/normal pair had sufficient coverage for variant calling (**S3 Table**). Using a combination of somatic variant calling algorithms and stringent filtering parameters, we identified 2,879 high-confidence coding and splice-site somatic variants (consisting of 2,214 nonsynonymous (1,405 SNVs, 809 indels), 92 splice-site, and 573 synonymous variants) (**S4 Table**). Combined, the 2,306 nonsynonymous and splice-site variants affected 1,968 protein-coding genes and averaged 153.7 variants per tumor (range 9-542 per tumor) (**S4** and **S5 Tables**). For the TCGA late-stage cohort (n=66), we extracted a total of 28,996 somatic coding and splice-site variants distributed among 10,504 protein-encoding genes (**S6** and **S7 Tables**). Using MutSigCV (v1.4), we identified a total of 14 unique late-stage SMGs (**Fig 1**), representing 6 SMGs (q-value ≤0.1) in the NHGRI (**Table 1**) and 12 SMGs in the TCGA late-stage EEC cohorts (**Table 2**).

**Table 1.** Statistically significantly mutated genes (q≤0.10) identified within the NHGRI cohort of 15 late-stage EEC exomes.

| Gene symbol | Gene name | Number and frequency (%) of NHGRI late-stage tumors with non-silent mutation(s)** | p-value | q-value |
| --- | --- | --- | --- | --- |
| *PTEN* | Phosphatase and tensin homolog | 13 (86.7%) | 2.11E-15 | 3.98E-11 |
| *ARID1A* | AT-rich interaction domain 1A | 11 (73.3%) | 1.27E-10 | 1.20E-06 |
| *RPL22* | Ribosomal protein L22 | 4 (26.7%) | 1.44E-07 | 9.07E-04 |
| *OR6C75* | Olfactory receptor family 6 subfamily C member 75 | 4 (26.7%) | 7.84E-06 | 3.70E-02 |
| *CTCF* | CCCTC-binding factor | 5 (33.3%) | 1.26E-05 | 4.74E-02 |
| *AP1S1* | Adaptor related protein complex 1 subunit sigma 1 | 3 (20.0%) | 2.13E-05 | 6.70E-02 |

\*\* Non-silent mutations consist of nonsynonymous and splice junction mutations

**Table 2.** Statistically significantly mutated genes (q≤0.10) identified among 66 late-stage TCGA EECs.

| Gene symbol | Gene name | ξ Number and frequency (%) of TCGA late-stage tumors with non-silent mutation(s) | p-value | q-value |
| --- | --- | --- | --- | --- |
| *ARID1A* | AT-rich interaction domain 1A | 29 (43.9%) | 0 | 0 |
| *PIK3R1* | Phosphoinositide-3-kinase regulatory subunit 1 | 21 (31.8%) | 5.55E-16 | 5.24E-12 |
| *PTEN* | Phosphatase and tensin homolog | 46 (69.7%) | 2.00E-15 | 1.26E-11 |
| *PIK3CA* | Phosphatidylinositol-4,5-bisphosphate 3-kinase catalytic subunit alpha | 25 (37.9%) | 7.27E-14 | 3.43E-10 |
| *TP53* | Tumor protein p53 | 19 (28.8%) | 3.07E-13 | 1.16E-09 |
| *KRAS* | KRAS proto-oncogene, GTPase | 18 (27.3%) | 7.22E-11 | 2.27E-07 |
| *RPL22* | Ribosomal protein L22 | 6 (9.1%) | 4.89E-08 | 1.08E-04 |
| *CTCF* | CCCTC-binding factor | 12 (18.2%) | 4.99E-08 | 1.08E-04 |
| **CTNNB1** | Catenin beta 1 | 18 (27.3%) | 5.16E-08 | 1.08E-04 |
| **PAX6** | Paired box 6 | 7 (10.6%) | 1.04E-06 | 1.96E-03 |
| **RNF43** | Ring finger protein 43 | 9 (13.6%) | 4.61E-06 | 7.91E-03 |
| **KLF3** | Kruppel like factor 3 | 7 (10.6%) | 6.34E-06 | 9.96E-03 |

ξ Data were extracted from previously published TCGA data
\*\* Non-silent mutations consist of nonsynonymous and splice junction mutations

**Figure 1.**
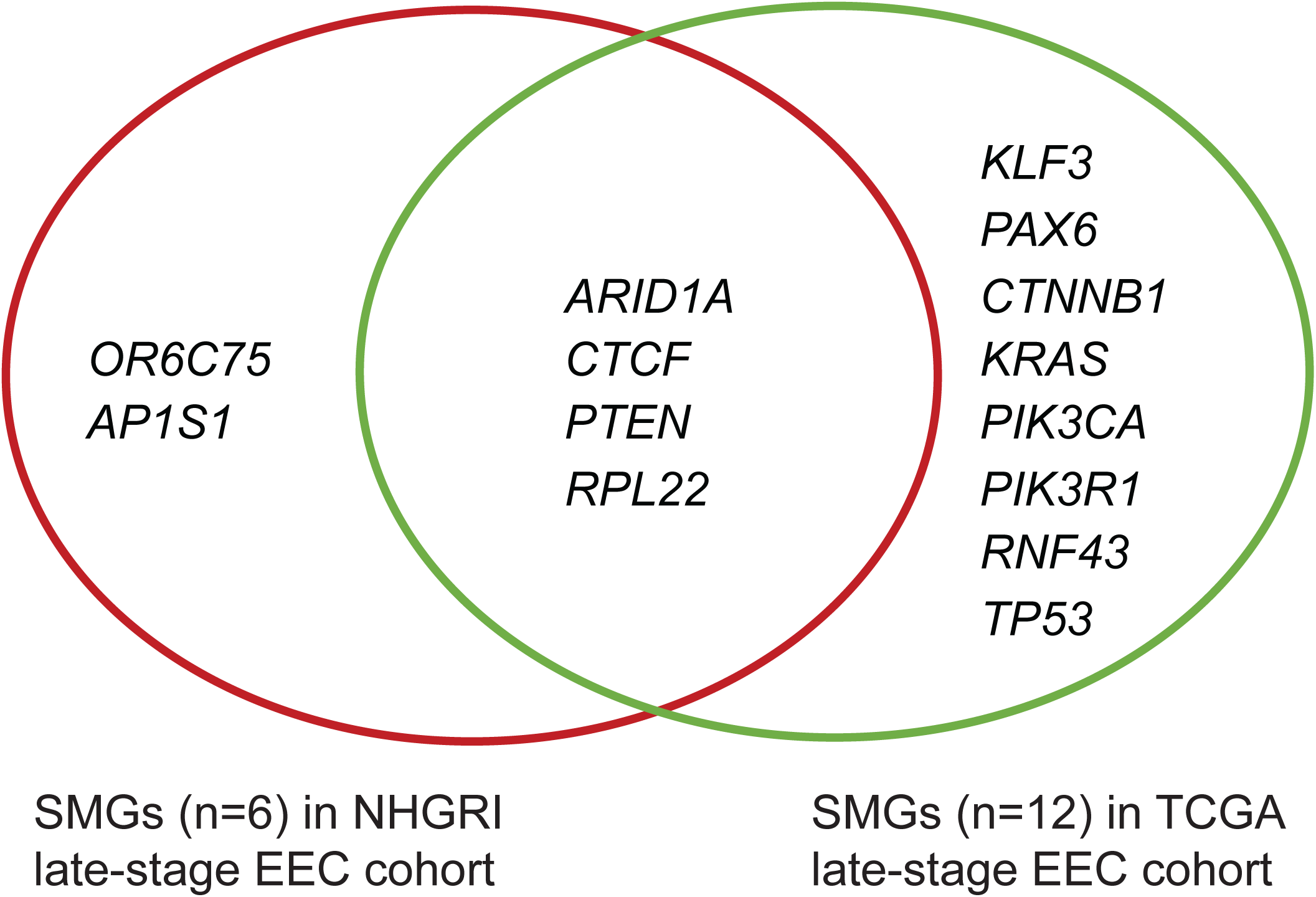
Statistically significantly mutated genes (SMGs) in late-stage and early-stage EEC cohorts. Venn diagram showing unique and shared SMGs identified by MutSigCV (v1.4) analysis of 15 NHGRI late-stage EECs and 66 TCGA late-stage EECs.

### *KLF3* and *PAX6* are SMGs in late-stage but not early-stage EEC

To test whether each of the 14 late-stage SMGs are also statistically significantly mutated in the TCGA early-stage EECs (n=270), we first estimated MutSigCV’s power to detect genes as significantly mutated in the early-stage cohort. Estimating power using a binomial model as described in [35], we determined that the data from 270 tumors, when tested on 14 genes, yields >95% power to detect genes as significantly mutated across a wide range of background mutation rates when at least 10% of the 270 tumors are mutated in that gene (**S1 Figure**). Next, we obtained somatic variants for the cohort of non-ultramutated TCGA early-stage EECs; there were 162,763 somatic coding- and splice-site variants affecting 17,435 protein-encoding genes (**S8** and **S9 Tables**). To determine whether any of the 14 late-stage SMGs were significantly mutated in this dataset, p-values for all somatically mutated genes in early-stage tumors were calculated and used to determine q-values adjusting for 14 tests (reflecting the 14 late-stage SMGs queried) using the Benjamini-Hochberg procedure [36] (**Table 3**). Results showed that 12 of 14 late-stage SMGs were statistically significantly mutated (q-value <0.1) in early-stage EECs whereas two late-stage SMGs, *KLF3* (Krüppel Like Factor 3) and *PAX6* (Paired Box 6), were not (**Table 3**). Somatic mutations were more frequent among late-stage tumors than early-stage tumors for both *KLF3* (10.6% (7 of 66) late-stage vs 4.8% (13 of 270) early-stage) and *PAX6* (10.6% (7 of 66) late-stage vs 1.9% (5 of 270) early-stage) (**Table 4**).

**Table 3.** *PAX6* and *KLF3* are the only late-stage EEC SMGs (q-value ≤0.1) that are not statistically significantly mutated in early-stage EEC.

| Gene symbol for 14 SMGs (q-value ≤0.1), identified in late-stage EEC cohorts | MutSigCV (1.4) p-value in early-stage EEC TCGA cohort | MutSigCV (1.4) q-value in early-stage EEC TCGA cohort corrected for 14 tests |
| --- | --- | --- |
| *AP1S1* | 0.00E+00 | 0.00E+00 |
| *ARID1A* | 0.00E+00 | 0.00E+00 |
| *CTNNB1* | 0.00E+00 | 0.00E+00 |
| *PIK3CA* | 0.00E+00 | 0.00E+00 |
| *PTEN* | 0.00E+00 | 0.00E+00 |
| *RNF43* | 0.00E+00 | 0.00E+00 |
| *RPL22* | 0.00E+00 | 0.00E+00 |
| *TP53* | 1.11E-15 | 1.94E-15 |
| *CTCF* | 2.78E-15 | 4.05E-15 |
| *PIK3R1* | 2.89E-15 | 4.05E-15 |
| *KRAS* | 6.88E-15 | 8.76E-15 |
| *OR6C75* | 1.89E-03 | 2.21E-03 |
| *PAX6* | 1.29E-01 | 1.39E-01 |
| *KLF3* | 9.99E-01 | 9.99E-01 |

**Table 4.** Frequency of non-silent *KLF3* and *PAX6* mutations in non-ultramutated EECs, according to molecular subgroup.

| ξTumor stage and molecular subgroup | ξ*KLF3* mutation frequency | ξ*PAX6* mutation frequency |
| --- | --- | --- |
| **All stages of EEC (n=336)** | **5.9% (20 of 336)** | **3.6% (12 of 336)** |
| MSI subgroup (n=141) | 14.2% (20 of 141) | 7.8% (11 of 141) |
| CN low subgroup (n=140) | 0% (0 of 140) | 0.7% (1 of 140) |
| CN high subgroup (n=55) | 0% (0 of 55) | 0% (0 of 55) |
| **Late-stage EECs (n=66)** | **10.6% (7 of 66)** | **10.6% (7 of 66)** |
| MSI subgroup (n=27) | 25.9% (7 of 27) | 25.9% (7 of 27) |
| CN low subgroup (n=21) | 0% (0 of 21) | 0% (0 of 21) |
| CN high subgroup (n=18) | 0% (0 of 18) | 0% (0 of 18) |
| **Early-stage EECs (n=270)** | **4.8% (13 of 270)** | **1.9% (5 of 270)** |
| MSI subgroup (n=114) | 11.4% (13 of 114) | 3.5% (4 of 114) |
| CN low subgroup (n=119) | 0% (0 of 119) | 0.8% (1 of 119) |
| CN high subgroup (n=37) | 0% (0 of 37) | 0% (0 of 37) |

ξ Data were extracted from previously published TCGA data

### Late-stage-specific EEC SMG (*KLF3* and *PAX6*) mutations occur in MSI-hypermutated EEC and are predicted to affect protein function

For the TCGA cohorts, we evaluated the distribution of *KLF3* and *PAX6* mutations across the MSI-hypermutated (n=141 cases), CN-low (n=140 cases), and CN-high (n=55 cases) molecular subgroups (**Table 4**). *KLF3* mutations occurred exclusively in the MSI-hypermutated subgroup at an overall frequency of 14.2% (20 of 141 cases), which was significantly higher than the occurrence of *KLF3* mutations among the combined CN-high and CN-low subgroups (0 of 195 cases) (p-value < 0.0001 2-tailed Fisher’s exact test). Within the MSI-hypermutated subgroup, *KLF3* was mutated in 25.9% (7 of 27) of late-stage tumors *versus* 11.4% (13 of 114) of early-stage tumors. There were no statistically significant differences in *KLF3* mutation frequency according to tumor grade; mutations were present in 14.2% of grade 1 (4 of 28), 8.1% of grade 2 (3 of 37), and 13.2% of grade 3 (11 of 83) MSI tumors (**S10 Table**).

All but one (11 of 12) of *PAX6* mutations were in the MSI subgroup; the *PAX6*^X306_splice^ mutation was present in a CN-low tumor (**Table 4**). The higher frequency of *PAX6* mutations in the MSI-hypermutated subgroup compared to other subgroups was statistically significant (p-value = 0.0004, 2-tailed Fisher’s exact test). Within the MSI-hypermutated subgroup, *PAX6* was mutated in 7.8% (11 of 141) of tumors; mutations in late-stage tumors were more frequent compared to early-stage tumors (25.9% (7 of 27) *versus* 3.5% (4 of 114)). There was no significant difference in the frequency of *PAX6* mutations between tumors of differing grade; *PAX6* mutations were present in 3.6% of grade 1 (1 of 28), 13.5% of grade 2 (5 of 37) and 7.9% of grade 3 (6 of 76) MSI-hypermutated tumors (**S10 Table**). We observed no statistically significant differences in *KLF3* or *PAX6* mutation frequencies between *POLE*/*POLD1*-mutated and *POLE*/*POLD1*-wildtype cases within the MSI-hypermutated subgroup (**S11 Table**).

A majority of *KLF3* and *PAX6* mutations were indels within homopolymer tracts, resulting in frameshifts; the KLF3^K106Nfs*21^, KLF3^P226Rfs*52^, KLF3^Q227Afs*37^, and PAX6^P375Hfs*7^ frameshift mutations were recurrent (**Fig 2**). Six of 21 (28.6%) *KLF3* mutations and 3 of 11 (27.3%) *PAX6* mutations were missense mutations; KLF3^R257W^, KLF3^R261G^ and PAX6^A33T^ were predicted to affect protein function by 3 of 4 *in silico* algorithms (**S12 Table**).

**Figure 2.**
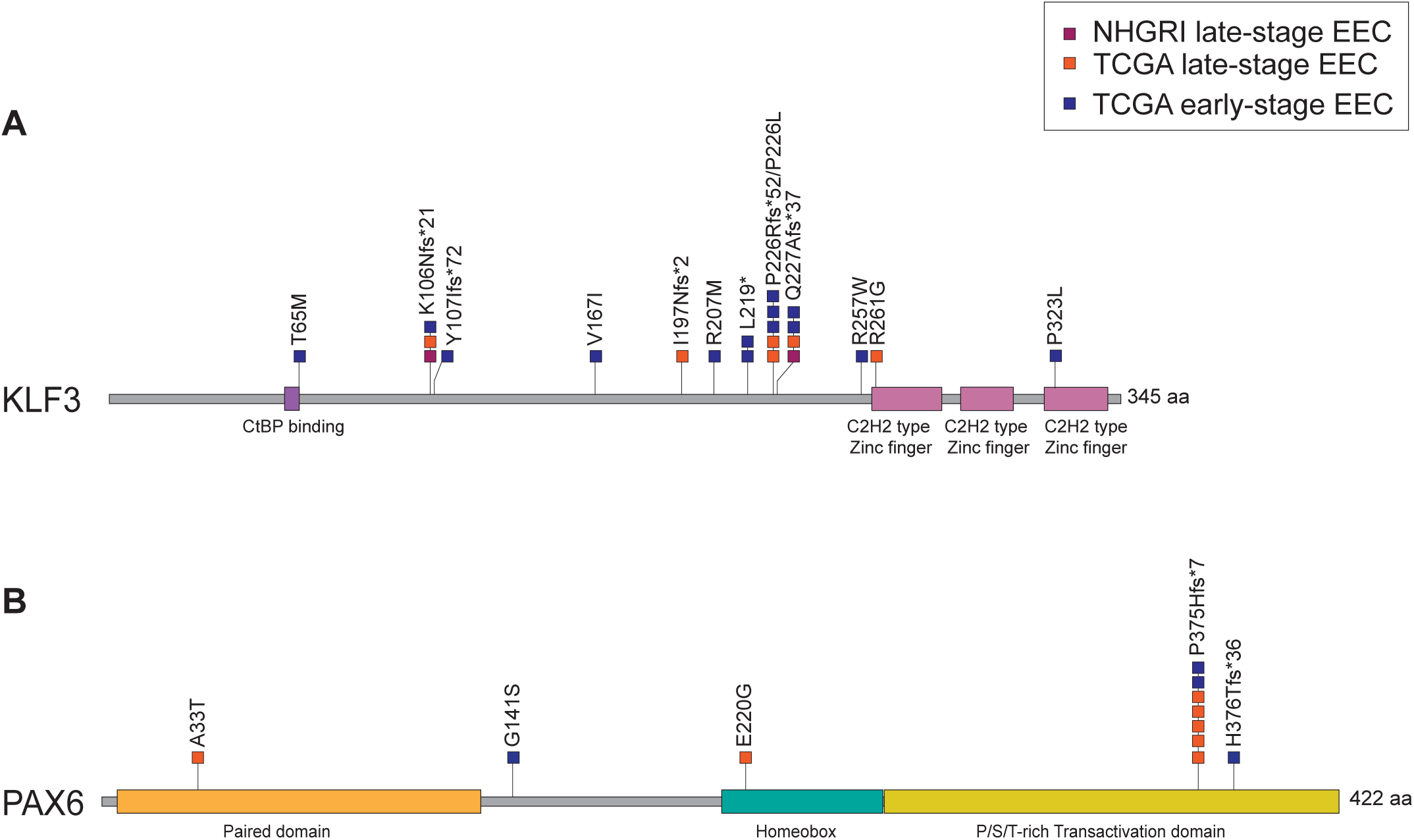
Spectrum of *KLF3* and *PAX6* somatic mutations in late-stage and early-stage non-ultramutated EECs. Lollipop plots showing the positions of somatic mutations in (A) KLF3 and (B) PAX6 relative to protein domains. Mutations in NHGRI and TCGA late-stage EEC (red and orange) cohorts and the TCGA early-stage (blue) EEC cohort are distinguished.

### Survival analysis

We utilized the cBioPortal for Cancer Genomics (https://www.cbioportal.org/) to query the relationship between patient survival and somatic mutation status of all 14 late-stage SMGs identified herein, applying a Bonferroni correction to account for multiple testing (672 tests). With respect to *KLF3* and *PAX6* in the MSI-hypermutated subgroup, no significant differences in overall survival (OS), progression-free survival (PFS), disease-free survival (DFS) or disease-specific survival (DSS) were observed between mutated and non-mutated tumors when all stages were combined or when early- and late-stage tumors were considered separately (**S13 Table**). For the remaining 12 SMGs, there were no statistically significant differences in survival for any stage or molecular subgrouping (data not shown).

## Discussion

The mutational landscape of EEC was reported by TCGA in an initial 2013 study and a subsequent “pan-gyn” study which included the 2013 EEC cohort and additional cases. Both studies performed *in silico* annotation of SMGs, which represent candidate driver genes, in a stage-agnostic manner. However, cancer genomes are dynamic and the mutational repertoire of tumors can evolve during progression and metastasis [37]. Recent comparisons of primary and metastatic endometrial cancer genomes have demonstrated diverge sease, raising the possibility that *KLF3* and *PAX6* mutations undergo positive selection during tumor progression.

*KLF3* encodes a zinc nce in their mutational landscapes [38-40]. But exome-wide comparisons of late-stage and early-stage primary tumors are lacking. Here, our stage-specific analysis of TCGA mutation data for non-ultramutated EECs showed that *KLF3* and *PAX6* are SMGs in late-stage (III/IV) but not early-stage (I/II) di finger transcription factor with roles in adipogenesis, erythroid maturation, B-cell differentiation, and cardiovascular development (reviewed in [41]). The encoded protein includes an N-terminal CtBP-binding motif, three C-terminal Cys2His2 zinc finger domains, and a primary phosphorylation site at serine-249 that is important for DNA binding and enhancing transcriptional repression [41]. In our analysis of NHGRI EEC exomes and TCGA mutation data, the majority of *KLF3* mutations, including three mutation hotspots, were frameshift mutations that occur N-terminal to the zinc finger domains and to serine-249. It is likely that these mutations result in nonsense-mediated decay and haploinsufficiency because the associated premature stop codons are located more than 50-55 nucleotides upstream of the final exon-exon junction [42]. In addition, *in silico* analyses predicted deleterious effects for the KLF3^R257W^ and KLF3^R261G^ missense mutants that occur in EEC; KLF3^R257W^ also occurs somatically in 2 colorectal cancers (1 MSI-high/CIMP (CpG island methylator phenotype)-low; 1 CIN (chromosome instability)-subgroup) [43, 44].

The fact that *KLF3* mutations in EEC occur predominantly at homopolymer tracts, were restricted to the MSI-hypermutated EEC subgroup, and are more frequently mutated in late-stage than early-stage MSI-hypermutated tumors (25.9% versus 11.4%, respectively), indicate that *KLF3* is an MSI target gene that may be involved in the etiology and progression of a subset of hypermutated EECs. Consistent with the idea that *KLF3* is an MSI target gene, frameshift mutations at codons 106 and 227, which are recurrent in MSI-EECs, are also recurrent in the colorectal MSI-colorectal and MSI-stomach TCGA molecular subgroups [27, 28, 44, 45].

Studies in other tumor types have reported *KLF3* alterations as adverse prognosticators. For example, decreased *KLF3* expression in colorectal and cervical cancers is associated with lymph node positivity and poorer outcomes [46, 47]. Conflicting data exist regarding the occurrence and effects of reduced *KLF3* levels in lung cancer. However, one study reported lower levels of *KLF3* mRNA and protein expression in lung adenocarcinomas compared with adjacent normal tissues and more frequent loss of KLF3 expression in late-versus early-stage disease [48]. Although we found *KLF3* is a late-stage-specific SMG in EEC, there was no significant association between *KLF3* mutation status and survival for EEC patients, possibly reflecting tissue-specific differences in *KLF3* association with outcome, and/or outcome differences between mutation and reduced expression of *KLF3*.

The second late-stage-specific SMG identified in our study was *PAX6* (paired box protein Pax-6). PAX6 encodes a highly conserved paired box transcription factor that includes paired box and homeobox DNA-binding domains and a C-terminal transactivation domain (TAD); the final 40 residues of the TAD influence homeobox-DNA binding [49]. PAX6 has important roles in the development of several tissue types, including the eye (reviewed in [50]). Inherited and *de novo* nonsense and frameshift mutations in *PAX6* cause the autosomal dominant eye disorder aniridia 1, whereas germline missense mutations are associated with attenuated ocular phenotypes [51]. Dysregulation of *PAX6* expression has been implicated in a variety of human cancers, resulting in tumor suppressive or oncogenic phenotypes depending on the cellular context [52-64]. A recent study reported a potential role for epigenetic silencing of *PAX6* in EC progression based on hypermethylation of *PAX6* in primary EC *versus* endometrial hyperplasia, and in metastatic EC *versus* primary EC [65]. Our analysis of TCGA mutation data found that *PAX6* mutations almost exclusively occur in MSI-hypermutated tumors. This observation, coupled with the fact that *PAX6* mutations were more frequent among late-stage than early-stage MSI-hypermutated tumors (25.9% *versus* 3.5%, respectively), raise the possibility that, like *KLF3* mutations, *PAX6* mutations may be pathogenic drivers of tumor progression in the context of MSI-hypermutated EECs.

Most *PAX6* mutations in TCGA MSI-hypermutated EECs were the recurrent *PAX6*^P375Hfs*7^ frameshift mutation in the transactivation domain [11, 13]. We predict that *PAX6*^P375Hfs*7^ and an adjacent *PAX6*^H376Tfs*36^ frameshift mutation encode truncated proteins with reduced transactivation capacity, because the associated premature stop codons are located within 50 nucleotides of the penultimate exon-exon junction and are located proximal to a synthetic nonsense mutation (PAX6^Q422X^) that exhibits reduced transactivation capacity *in vitro* [66]. Moreover, the fact that the PAX6^P375Q^ aniridia-associated missense mutation results in attenuated DNA binding affinity *in vitro* [66], raises the possibility that the recurrent PAX6^P375Hfs*7^ mutant also may have attenuated DNA binding. Similar to *KLF3* frameshift mutations, the PAX6^P375Hfs*7^ and PAX6^H376Tfs*36^ frameshift mutations in EEC both arise within a (C)_7_ homopolymer tract indicating that *PAX6* is an MSI target gene. Consistent with this idea is the fact that *PAX6* frameshift mutations originating at codon 375 and/or codon 376 are also recurrent in MSI-stomach cancer and MSI-colorectal carcinoma [27, 28, 45].

Compared to frameshift mutations, *PAX6* missense mutations are relatively rare in the non-ultramutated TCGA cohort, occurring in three cases. The PAX6^A33T^ EC-mutant occurs in the N-terminal paired box domain at a residue highly conserved across paired domains in Pax family members and other proteins and is predicted to impact function [67]. A different substitution at this residue (PAX6^A33P^) exhibits altered transactivation activity *in vitro* and is a germline variant associated with partial aniridia [67, 68]. The other two *PAX6* missense mutations in EC (PAX6^E220G^ and PAX6^G141S^) were not uniformly predicted to be functionally significant in our analysis and, to our knowledge, are not pathogenic variants for ocular phenotypes.

In conclusion, our findings indicate that *KLF3* and *PAX6* are candidate driver genes in a subset of late-stage hypermutated EECs and are MSI target genes. Despite sufficient power, neither *KLF3* nor *PAX6* were detected as candidate driver genes in early-stage EECs. To our knowledge, this is the first study to annotate *KLF3* and *PAX6* as late stage-specific SMGs in EEC. Our findings warrant future studies to independently validate the enrichment of *PAX6* and *KLF3* mutations in late-stage, MSI-hypermutated EECs and to determine the functional effects of recurrent frameshift mutations in these genes particularly in regard to phenotypic properties associated with tumor progression.

## Data Availability

Exome sequencing data for the NHGRI tumor-normal cohort have been deposited in dbGAP under controlled access

https://www.ncbi.nlm.nih.gov/projects/gap/cgi-bin/study.cgi?study_id=phs001153.v1.p1

