## Supplementary material for "*KLF3* and *PAX6* are candidate driver genes in late-stage, MSI-hypermutated endometrioid endometrial carcinomas": S1 Figure

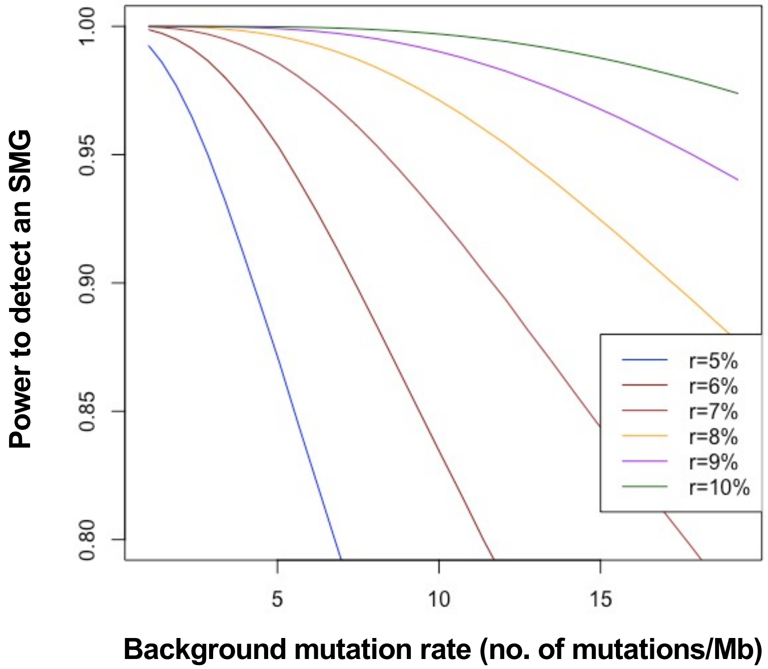


**S1 Figure.** Power to detect significantly mutated genes (SMGs) in early-stage tumors. Curves show statistical power for different percentages (r) of tumors that are somatically mutated. Calculations were performed as described in the text, assuming 270 tumors and 14 gene tests completed.
